# Selection bias in Mendelian randomization studies with adjustment for medication use

**DOI:** 10.64898/2026.09.16.26363225

**Authors:** Joy Shi, Sonja A. Swanson, Elizabeth W. Diemer, Hanna Gerlovin, Daniel C. Posner, Peter W.F. Wilson, J. Michael Gaziano, Kelly Cho, Miguel A. Hernán, the VA Million Veteran Program

## Abstract

**Background:** Mendelian randomization (MR) studies often evaluate exposures such as LDL cholesterol (LDL-C). The widespread use of lipid lowering medications complicates the interpretation and the validity of MR estimates.

**Methods:** We describe two causal estimands in populations with medication use: a lifetime effect, and a lifetime effect under no medication use. In simulations, we compared the common approach of excluding medication users with estimation based on inverse probability (IP) weighting to adjust for medication use. We applied both approaches to a MR analysis of LDL-C and coronary artery disease in the Million Veteran Program (MVP), a large prospective cohort of U.S. veterans with linked electronic health record and genetic data.

**Results:** In simulations, MR analyses that did not adjust for medication use estimated a lifetime effect that reflected a valid estimate of a total effect that included both the harms of higher LDL-C and the benefits of statins. To estimate the effect under no medication use, excluding statin users introduced selection bias. Alternatively, IP weighting could address bias from incident statin users, but could not address bias related to prevalent medication use. Estimates from MVP data varied considerably, reflecting the importance of these analytic choices.

**Conclusions:** Different medication adjustment strategies in MR studies implicitly target different causal estimands and are subject to distinct biases. Transparent analytical choices and careful interpretation are essential for informative MR results in the context of widespread medication use.

## Introduction

Mendelian randomization (MR) is an application of instrumental variable (IV) analysis in which genetic variants are proposed as instruments to estimate causal effects of non-genetic exposures. Many MR studies are interested in exposures, such as LDL-C or blood pressure, whose values are substantially affected by commonly used medications (e.g., statins and other lipid medications, antihypertensives) in the study population.^1–5^ Medication use makes the findings of such MR studies challenging to interpret. One problem is that the genetic variants become weaker instruments. For example, if many individuals in the study already use statin therapy, which lowers their LDL-C levels, the relation between genetic variants and LDL-C may be greatly attenuated. Also, when the interest lies in a potential effect of LDL-C in the absence of medication use, adjusting for medication use will generally introduce selection bias.^6^

Here, we argue that attempts to adjust for medication use cannot, in general, remove this selection bias in MR analyses. We evaluate the bias of different adjustment methods using, as an example, the effect of LDL-C and coronary artery disease. Our paper is structured as follows. We begin by describing two causal estimands in MR studies with time-varying exposures and medication use: a lifetime effect and a lifetime effect under no medication use. Next, when targeting the latter estimand, we outline different approaches to adjust for medication use. We then use simulated data to compare the direction and magnitude of bias introduced by each approach. Last, we apply these approaches to an MR analysis of LDL-C and coronary artery disease using data from the Million Veteran Program (MVP) to highlight practical considerations for each adjustment method.

### Causal estimands in MR studies with time-varying exposures and medication use

The causal directed acyclic graph (DAG) in Figure 1 depicts the proposed genetic instrument (*Z*), the time-varying exposure (*A_k_*), time-varying medication use (*M_k_*), indicators for a failure-time outcome (*Y_k_*_+1_), exposure-outcome confounders (*U*), and medication use-outcome confounders (*W*). In Figure 1, the proposed instrument *Z* satisfies the three instrumental conditions: (1) it is associated with the exposure; (2) it does not affect the outcome except through its possible effect on the exposure; and (3) it does not share causes, or other sources of lack of exchangeability, with the outcome.^7^ For simplicity, we show only two time points, *k* = 0 and *k* = 1, but our discussion applies to more general settings with multiple time points.

**Figure 1.**
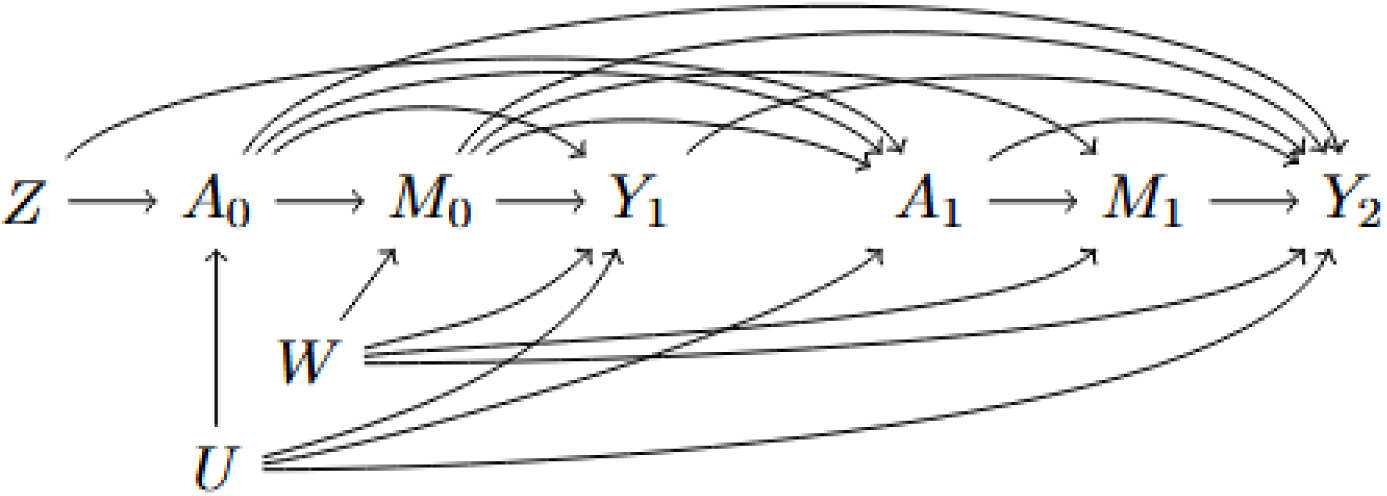
Causal diagrams for a Mendelian randomization analysis for a time-varying exposure affected by medication use. Variables included are a genetic instrument *Z*, time-varying exposure *A_k_*, time-varying medication use *M_k_*, outcome *Y_k_*, exposure-outcome confounders *U*, and medication use-outcome confounders *W*.

As an example, suppose *A_k_* represents LDL-C, *M_k_* is an indicator of statin use, and *Y_k_*_+1_ is an indicator of coronary artery disease at time *k* + 1. Two possible effects of interest are the *lifetime effect of LDL-C* and the *lifetime effect of LDL-C under no medication use*, which we now describe (see Supplement 1 for formal definitions).

The *lifetime effect* of LDL-C can be interpreted as a contrast between the disease risks under (i) a hypothetical intervention that increases the natural value of LDL-C at each time *k* by a fixed quantity over the lifetime (starting, perhaps, from the gestational age when LDL-C starts to circulate in the blood in humans) without affecting the risk of coronary artery disease via other mechanisms not mediated through LDL-C, and (ii) the observed value of LDL-C.^7,8^ The natural value of LDL-C at time *k* is the value of LDL-C that would have been observed at *k* had the hypothetical intervention described above been discontinued right before *k* . The observed value of LDL-C at time *k* is the natural value in our world in which the hypothetical intervention was never implemented.

Note that the natural value of LDL-C has changed since statins became available because, in contemporaneous populations, high levels of LDL-C often trigger a prescription for statins, which lowers subsequent LDL-C levels. That is, the interpretation of the lifetime effect of LDL-C generally depends on the prevalence of statin use, which complicates comparisons across studies in different regions and times.

An alternative interpretation of the lifetime effect is a contrast between the disease risks under (iii) a hypothetical intervention that manages to keep LDL-C at a fixed value (e.g., 100 mg/dL) over the lifetime and (iv) a hypothetical intervention that manages to keep LDL-C at another fixed value (e.g., 139 mg/dL) over the lifetime, with both interventions (iii) and (iv) not affecting the risk of coronary artery disease via other mechanisms not mediated through LDL-C. This contrast eliminates the dependence of the effect on the potentially changing magnitude of the natural value of LDL-C, but the intensity of the hypothetical interventions (iii) and (iv) will depend on whether the person is receiving statins, and the effects of these hypothetical interventions may be partially mediated by future statin use. Therefore, this estimand would not be particularly relevant if investigators were interested in an effect of LDL-C that is independent of statin use.

Now let us consider the *lifetime effect under no medication use*, which we will define as the contrast of disease risks under expanded hypothetical interventions (iii) and (iv) that include a concurrent intervention to prevent statin use. (An alternative interpretation of this effect would be based on expanded interventions (i) and (ii), but we will not consider it here.) In this paper, we focus on both the lifetime effect comparing interventions (iii) and (iv) (hereafter referred to simply as “lifetime effect”), and the lifetime effect under no medication use, the latter being arguably the one targeted by MR analyses that adjust for medication use.

### Estimation procedures

Most MR studies for time-varying exposures are based on IV estimation via two-stage least squares (TSLS) using a single measure of the exposure and a binary end-of-follow-up outcome, and require linearity assumptions. This approach can correctly estimate the total lifetime effect of LDL-C (including through statin use) if *Z* satisfies the three instrumental conditions, as in Figure 1, and certain homogeneity conditions, including a constant gene-exposure association over time, hold.^7–9^ To relax the assumption of a constant gene-exposure association, it is necessary to use IV approaches that incorporate repeated exposure measures,^9^ such as g-estimation of structural nested cumulative failure time models (SCNFTM), which also allow for modeling the outcome as a failure time outcome.^10^ Regardless of the IV approach, the strength of the association between the genetic instrument *Z* and the exposure *A_k_* is weakened when earlier exposure levels trigger medication use that decreases later exposure levels, and weak instruments are a well-known threat to the validity of IV analyses.^7,11,12^

To estimate the lifetime effect of LDL-C under no medication use, investigators need to be able to jointly identify the effect of two hypothetical interventions: one that changes the value of LDL-C and one that prevents individuals from receiving statin therapy. This can be attempted by applying IV estimation (with the same caveats described in the previous paragraph) to a pseudo-population generated by inverse-probability (IP) weighting in which no individual receives statins. If follow-up in the MR study began before anyone has yet started statins, the pseudo-population is generated by censoring individuals if/when they start using statins and assigning the yet uncensored individuals to a time-varying nonstabilized IP weight whose denominator is, informally, the probability of remaining a non-statin user conditional on all time-fixed and time-varying confounders of the effect of statins (e.g., variables *W* and *A_k_* in Figure 1). Note that simply restricting the analysis to individuals who remain non-users throughout follow-up, as done in some MR studies, may introduce selection bias because statin use (*M_k_*) is a descendant of the collider LDL-C (*A_k_*) along the path *Z* → *A_k_* ← *U* → *Y*.^6,13^

However, MR studies often include participants who were already using statins at the start of follow-up. The inclusion of these prevalent users in the analysis may introduce bias (because they are, by definition, survivors of earlier statin initiation) that cannot be generally addressed by IP weighting.^14^ For example, in Figure 1, when follow-up starts at time 1 and prevalent users are included (i.e., no restriction on *M*_0_), restricting the analysis to individuals free of the outcome at time 1 (i.e., *Y*_1_ = 0) results in selection bias for statin use through the path *M*_1_ ← *M*_0_ → *Y*_1_ ← *U* → *Y*_2_. The exclusion of prevalent users from the analysis may also introduce selection bias due to conditioning on a collider (via the path *Z* → *A_k_* ← *U* → *Y* described above). Thus, in the presence of prevalent users, there is an inherent bias trade-off in MR analyses that target the lifetime effect of exposure under no medication. We explored this trade-off first using simulated data and then using real data from the Million Veteran Program.

## Simulated data

### Data-generating mechanisms

We simulated 500 longitudinal datasets of 50,000 individuals each based on the causal DAG in Figure 1. For each individual, we simulated up to 10 time points ranging from *k* = 0 (age 30) to *k* = 9 (age 75) or until the outcome occurred. The data generating mechanism used parametric models as described in Supplement 2. We considered two scenarios:

1. Null effect: Neither LDL-C nor statin use had a direct effect on coronary artery disease (i.e., in Figure 1, there are no arrows from *A_k_* to *Y_t_* and *M_k_* to *Y_t_* for all *k* < *t*). In this scenario, the lifetime effects of LDL-C with and without intervention on statins are equal and null.
2. Non-null effect: LDL-C had a direct harmful effect, while statin use had a direct protective effect. In this scenario, the lifetime effects of LDL-C with and without intervention on statins are different and non-null.

In both scenarios, the magnitude of the gene-exposure association changed over time. In the second scenario, the effect of LDL-C on coronary artery disease risk did not vary by statin use (i.e., a LDL-C level of 100 mg/dL contributed equally to disease risk on or off statins).

### Data analysis

In each simulated dataset, we estimated the risk ratio for an increase in LDL-C from 100 mg/dL to 139 mg/dL via both (1) two-stage least squares (TSLS), and (2) g-estimation of structural nested cumulative failure time models (SNCFTMs). We conducted separate analyses with follow-up starting at age 30 (i.e., using all 10 simulated time points) where all participants are non- or new statin users, and at ages 45 and 60 (i.e., using simulated data from the 4^th^ and 7^th^ time points) where some participants may be prevalent users at entry. We also conducted analyses with staggered entry, where participants began follow-up at varying ages randomly drawn from a normal distribution (mean 52.5, SD 6.25 years), to reflect the typical staggered entry observed in MR studies.

We obtained risk ratio estimates for the two estimands described above: a lifetime effect, based on analyses of all participants with no exclusions, and a lifetime effect under no medication use. For the latter, we considered three approaches: (1) exclusion of prevalent users (i.e., users before the start of follow-up); (2) exclusion of prevalent and incident users (i.e., users before and after the start of follow-up); and (3) exclusion of prevalent users plus IP weighting to adjust for confounders of the effect of statins during follow-up. For each analysis, we computed the mean and the 2.5^th^ and 97.5^th^ percentiles of the risk ratio estimates across the 500 samples. For the scenario with non-null effect, we estimated the true effects via the plug-in g-formula with the correct parametric models that included the unmeasured variables *U*. Additional details on the statistical analysis are available in Supplement 3.^10^

### Simulation results

Figure 2 shows the results for scenarios 1 (null effects) and 2 (non-null effects). Under scenario 1, estimates were unbiased for the lifetime effect when the analysis included all participants, regardless of statin use. When targeting the lifetime effect under no medication use, excluding statins users (prevalent or incident) introduced bias, with increasing bias as the age (and therefore the proportion of prevalent users) increased. As expected, IP weighting could correct the bias due to excluding incident users but not the bias due to excluding prevalent users. Results were similar for both IV methods since data were simulated under the null.

**Figure 2.**
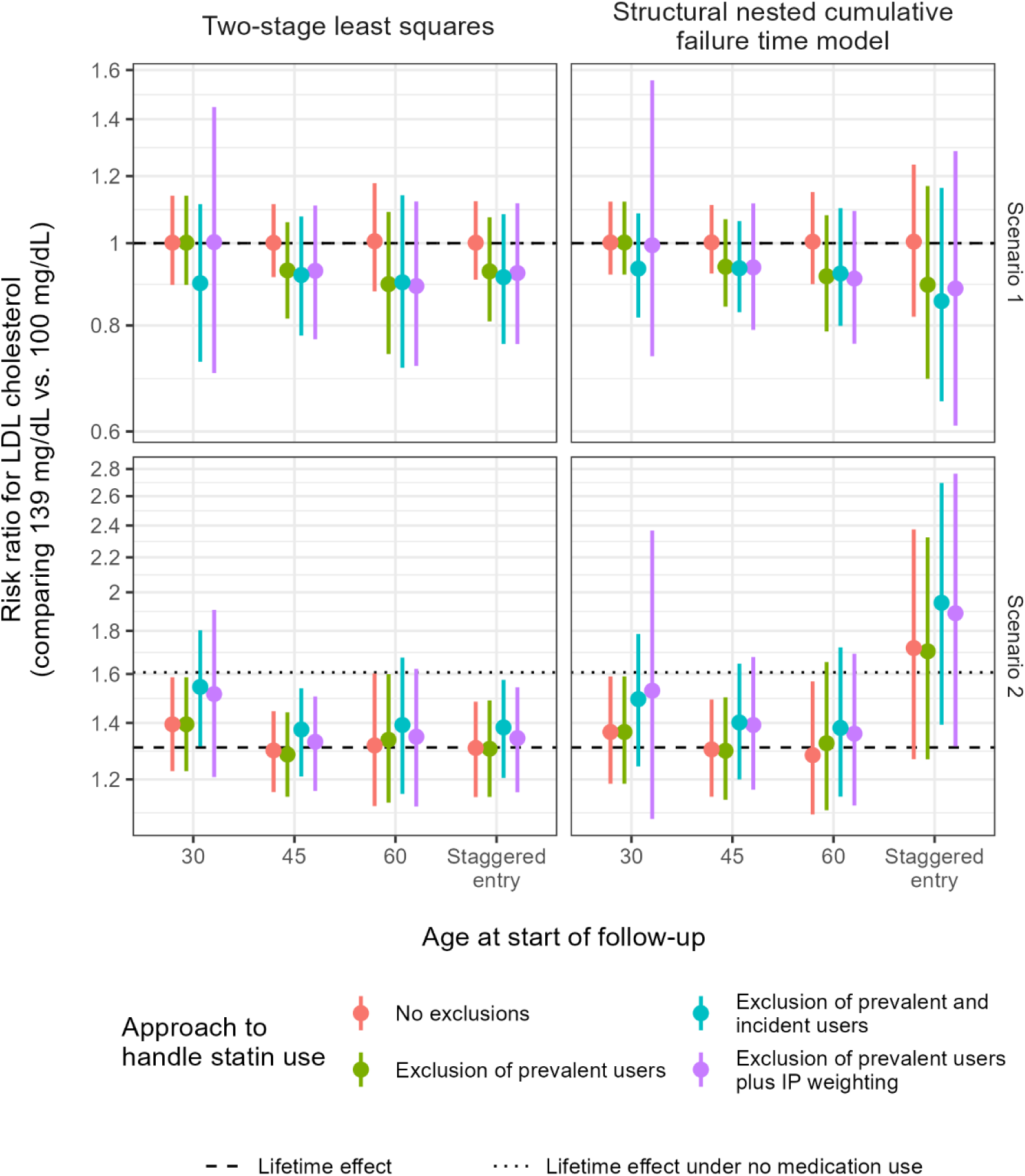
Risk ratio estimates under two IV methods, summarized across 500 iterations (by mean and 2.5th and 97.5th percentiles). In scenario 1, the effect of LDL cholesterol and statins is null (both the dotted and dashed lines are at the null). In scenario 2, LDL cholesterol had a direct harmful effect, while statins had a direct protective effect.

In scenario 2, estimates obtained under no exclusions were similar to the lifetime effect of LDL-C when follow-up began at age 30. When follow-up began at older ages, a separate selection bias arose from excluding participants with prevalent outcomes at entry (see Supplement 4).^15^ This bias was not observed in scenario 1, as it does not occur under the null. When targeting the lifetime effect under no medication use, the exclusion of statin users (prevalent or incident) biased the estimates upwards (opposite to the downward bias observed in scenario 1) due to new biasing pathways introduced by the direct effects of both LDL-C and statin use. The IP weighted SNCFTM estimate at age 30 was closest to the true lifetime effect under no medication use, but diverged as the age and proportion of prevalent statin users increased at the start of follow-up. Similar patterns were observed with the TSLS approach; however, its estimates were further from the true lifetime effect under no medication use, likely because it cannot account for changes in the gene-exposure association over time.

## Real data from the Million Veteran Program

### Study population

Study participants were from the Million Veteran Program (MVP), an ongoing observational prospective cohort study and biobank of active users of the Veterans Health Administration (VHA) health care system.^16^ Enrolment began in 2011, which entailed the collection of questionnaire data, blood samples and consent to data linkage with medical records and administrative health data from participants. All participants signed informed consent, and the study protocol was approved by the Veterans Affairs Central Institutional Review Board.^16^

For this study, eligible participants were MVP enrollees of European ancestry, aged 18 years or older between January 1, 2002 and December 31, 2019, with genotype data, no history of coronary artery disease or contraindications to lipid-lowering therapies (myopathy, chronic kidney disease or end stage renal disease, chronic liver disease, or abnormal liver function), at least one primary care visit in the past year, and documented body mass index (BMI), smoking status and LDL-C in the past year. Follow-up began when all eligibility criteria were first met (which could occur before or after MVP enrolment) and continued until the development of coronary artery disease, loss to follow-up (defined as ≥2 years from last recorded LDL-C measurement), 10 years after start of follow-up, or the administrative end of follow-up (December 31, 2020), whichever occurred first.

### Data collection

We used survey and electronic health record data from MVP core release v21.1, and genetic data from the MVP Release 4 dataset, for which genetic quality control methods have been previously described.^17^ For the proposed instrument, we constructed a weighted allele score based on 14 genetic variants in *HMGCR, PCSK9*, and *LDLR*, weighted by the magnitude of their association with LDL-C in the Global Lipids Genetics Consortium (GLGC).^18^

Longitudinal LDL-C measurements were obtained from outpatient blood specimens and identified using records from the Lab Chemistry domain in the VA’s corporate data warehouse (CDW) linked to MVP participants.^19,20^ LDL-C was estimated using the Friedewald formula and both fasting and non-fasting data were included in the analyses.^21^ The primary outcome of nonfatal or fatal coronary artery disease events (which included myocardial infarction, atherosclerosis, and undergoing angioplasty or revascularization procedures) was identified using records in the Outpatient and Inpatient domains in CDW, Centers for Medicare and Medicaid Services (CMS), and the National Death Index using a previously identified set of diagnosis and procedure codes (ICD-9, ICD-10 and CPT).^22,23^ Data on lipid-lowering therapies, including statins, ezetimibe, and bile acid sequestrants, were obtained from the Medications domain.^24^ Additional information on these measures and other covariates is provided in Table S1.

### Statistical analysis

As in our simulations, we estimated the lifetime effect of LDL-C without exclusions for lipid-lowering therapy use and the lifetime effect under no medication use using three approaches. For the IP weighting approach, nonstabilized time-varying weights were estimated by fitting pooled logistic regression models using 3-month discrete time intervals, with covariates including calendar time, age, sex, weighted allele score (i.e., the proposed instrument), education level, BMI, smoking status, lipid levels (LDL-C, HDL cholesterol, triglycerides, total cholesterol), systolic blood pressure, hospitalization in the past year, number of primary care visits in the past year, history of relevant comorbidities (myopathy, chronic liver disease, any vascular-related disease, diabetes, chronic obstructive pulmonary disease, depression, hypothyroidism, osteoporosis), and use of certain medications (antianginals, anticoagulants, antihypertensives and beta blockers).

We estimated 10-year risk ratios for an increase in LDL-C from 100 mg/dL to 139 mg/dL using TSLS and g-estimation of SNCFTM. All analyses adjusted for age, sex and ten genetic principal components of ancestry. To account for loss to follow-up, nonstabilized IP weights for censoring were constructed using calendar time, age, sex, and LDL-C. The final IP weights were derived from the IP weights for censoring, or their product with the IP weights for statin initiation when appropriate, and were truncated at the 99^th^ percentile. Confidence intervals were obtained using 500 bootstrap samples. Additional analyses were conducted by restricting to individuals in certain age groups (<60, 60 to <70) at the beginning of follow-up, where the same participant could contribute to multiple age group-specific analyses if eligibility criteria were met across multiple ages.

## Results

After applying the eligibility criteria, 327,063 MVP enrollees were included (Figure S2). At the time the eligibility criteria were met, participants had a mean age of 55.4 years, 91% were male, 28.8% had used a lipid-lowering therapy in the past year (Table 1).

**Table 1.** Characteristics at time of first eligibility for MVP participants included in MR analyses of LDL-C and coronary artery disease.

| **Characteristic** | **Summary statistic**<br>**(n = 327,063)** |
| --- | --- |
| Age (years) |  |
| Mean ± SD | 55.4 ± 13.1 |
| Median (range) | 57.6 (18.6, 102.1) |
| Calendar year, median (range) | 2008 (2002, 2019) |
| Sex, n (%) |  |
| Male | 297,952 (91.1) |
| Female | 29,111 (8.9) |
| BMI (kg/m<sup>2</sup>), mean ± SD | 29.9 ± 5.7 |
| Smoking status, n (%) |  |
| Never smoker | 118,777 (36.3) |
| Current smoker | 105,408 (32.2) |
| Former smoker | 102,878 (31.5) |
| Lipids (mg/dL), mean ± SD | |
| Total cholesterol | 192.1 ± 40.5 |
| Triglycerides | 161.1 ± 112.1 |
| HDL cholesterol | 45.4 ± 13.7 |
| LDL cholesterol | 116.7 ± 35.1 |
| Systolic blood pressure (mmHg), mean ± SD | 130.9 ± 15.8 |
| Hospitalized in the past 12 months, n (%) | 24,356 (7.4) |
| Number of primary care visits in the past 12 months, n (%) |  |
| 1 | 123,349 (37.7) |
| 2 | 88,083 (26.9) |
| 3 | 49,836 (15.2) |
| 4 | 27,175 (8.3) |
| $\geq 5$ | 38,620 (11.8) |
| History of comorbidities |  |
| Other vascular-related diseases | 14,877 (4.5) |
| Diabetes | 45,343 (13.9) |
| Chronic obstructive pulmonary disease | 19,601 (6.0) |
| Depression | 60,228 (18.4) |
| Hypothyroidism | 14,224 (4.3) |
| Osteoporosis | 2,907 (0.9) |
| Use of a lipid-lowering therapy in the past year, n (%) | 61,794 (18.9) |
| Medication use at time of first eligibility, n (%) |  |
| Lipid-lowering therapy | 94,352 (28.8) |
| Antianginals | 1,227 (0.4) |
| Anticoagulants | 6,164 (1.9) |
| Antihypertensives | 13,180 (4.0) |
| Beta blockers | 38,243 (11.7) |

Figure 3 presents MR estimates corresponding to 10-year risk ratios comparing LDL-C levels of 139 mg/dL versus 100 mg/dL in relation to coronary artery disease. Estimates from analyses without exclusions were consistent across IV methods, with risk ratios of 1.09 (95% CI: 0.97, 1.22) using TSLS and 1.06 (95% CI: 0.96, 1.20) using SNCFTM (Table S2). Excluding prevalent users of lipid-lowering therapies yielded similar risk ratios. However, excluding both prevalent and incident users led to a large upward shift in estimates, especially with TSLS (RR = 1.51, 95% CI: 1.18, 1.96) versus SNCFTMs (RR = 1.11, 95% CI: 0.92, 1.94), although confidence intervals were wide due to the high proportion of users excluded (n = 198, 211; 60.6%). IP weighted estimates were consistently larger than those from analyses without exclusions or with only prevalent users excluded. These patterns were consistent across both IV estimation approaches.

**Figure 3.**
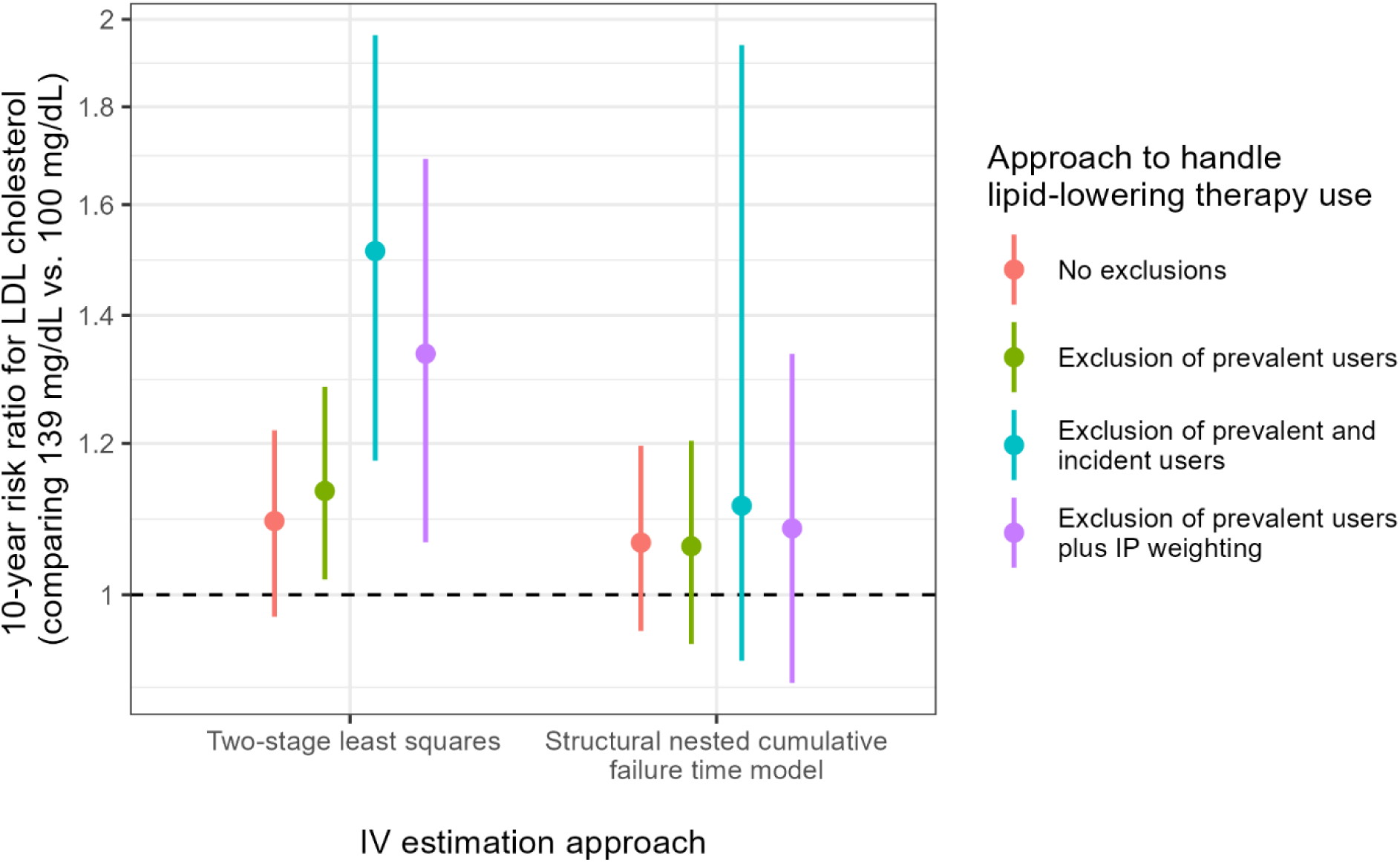
10-year risk ratio estimates and confidence intervals under two IV methods for the relationship between LDL cholesterol (139 mg/dL versus 100 mg/dL) and coronary artery disease in the Million Veteran Program.

In the age group-specific analyses (Table S2), the impact of different adjustment approaches on MR estimates was consistent with the overall findings, although estimates were larger in the <60 age group. Confidence intervals were especially wide for some analyses, such as those that excluded both prevalent and incident users. Additionally, the SNCFTM failed to converge in some bootstrap samples, which precluded the construction of valid confidence intervals.

## Discussion

We empirically evaluated MR analyses targeting either a lifetime effect of LDL-C or a lifetime effect of LDL-C under no medication use. In simulations, MR estimates of the lifetime effect were not biased, but MR estimates of the lifetime effect under no medication use based on excluding prevalent or incident statin users were biased. Although IP weighting could address the bias from excluding incident users, it could not address the bias from excluding prevalent users. In analyses of real data from MVP, MR effect estimates varied considerably by adjustment strategy, with the most extreme values observed when both prevalent and incident statin users were excluded.

Our analyses indicate that the lifetime effect under no medication targeted by MR analyses of time-varying exposures cannot be correctly estimated in the presence of prevalent users, regardless of whether the prevalent users are included^25–27^ or excluded^6^ from the analysis, because of selection bias. It is difficult to determine which approach yields less overall bias due to the presence of multiple and often opposing biasing pathways that depend on many factors, including the strength of the instrument, the prevalence of medication use, and the magnitude of effect of biomarker levels on medication use and vice versa.^6^ In the absence of prevalent users, IP weighting can mitigate the bias from excluding incident users, but only if all relevant factors related to statin initiation are adequately measured. Studies might avoid prevalent users by beginning follow-up at earlier ages. Unfortunately, because genetic cohorts used for MR, including MVP, typically enroll older participants, this is rarely possible. Another challenge is that statin use likely violates the homogeneity assumption required for identifying a lifetime effect in the population, as it modifies the gene-LDL-C association (by attenuating the association among statin users) and may also modify the effect of LDL-C on coronary artery disease risk (although our simulated data did not incorporate the latter). This homogeneity violation would introduce additional bias, which could be evaluated in future simulation studies.

We did not evaluate alternative MR estimators, such as those for two-sample MR, which often rely on summary statistics from datasets in which medication users have already been excluded. For example, the Global Lipids Genetics Consortium reports gene-lipid associations after excluding users of lipid-modifying therapies.^18^ Such exclusions can subsequently introduce selection bias into two-sample MR analyses. Furthermore, most robust MR methods cannot address this bias, as their alternative assumptions do not hold in this context.^6^ For example, some methods require only a subset of variants to be valid;^28^ however, this bias affects all variants because selection occurs downstream of the exposure.^6^ Similarly, approaches that assume independence between the bias and the gene-exposure association (e.g., MR-Egger) are also invalid because the biasing pathway involves the gene-exposure relationship.^6,29^

More fundamentally, the lifetime effects targeted by MR analyses of time-varying exposures affected by medication use are difficult to interpret. For example, the lifetime effect of LDL exposure is defined by hypothetical interventions that would sustain increases in LDL-C levels despite increasing statin use, which does not correspond to any known intervention and may not be physiologically attainable for all individuals. The already discredited analogy of MR studies as naturally occurring randomized trials is further challenged in studies where exposures are affected by medication use.^13,30^ The lifetime effect of LDL-C under no medication use is somewhat more interpretable, but still involves vague or hypothetical interventions, unlike the relatively well-defined interventions evaluated in trials.^31,32^ Furthermore, an intervention that completely withholds medication use may be unrealistic. A more realistic alternative would have been to consider dynamic strategies for medication use alongside the exposure of interest, such as allowing for statin initiation when LDL-C exceeds a certain threshold (e.g., ≥190 mg/dL). However, the causal estimands most commonly targeted in MR practice do not include dynamic strategies.

Medication use is often overlooked in MR studies. When researchers target effects under no medication use, we show different adjustment approaches for medication use can introduce bias of varying, and sometimes unpredictable, magnitude and direction. Our findings highlight the need to clearly define the causal question, including distinguishing between the lifetime effect and the effect under no medication use, and to select appropriate estimation strategies given the estimand of interest.

## Data Availability

Data used in this study cannot be shared publicly because of VA policies on data privacy and security and currently only available to approved VA researchers through a research merit review process with VA's Office of Research and Development (ORD).

## Acknowledgments

The authors thank Juan P. Casas, the VA Informatics and Computing Infrastructure (VINCI) and Genomic Information System for Integrative Science (GenISIS) support teams and the MVP Core Statistical Analysis team for their contributions to this study, as well as the Veterans who agreed to enroll in MVP. This publication does not represent the views of the Department of Veteran Affairs or the United States Government.

