## Supplemental Files for "Selection bias in Mendelian randomization studies with adjustment for medication use"

### Supplementary Materials

### Acknowledgement List: VA Million Veteran Program

**MVP Program Office**
- Sumitra Muralidhar, Ph.D., Program Director
  US Department of Veterans Affairs, 810 Vermont Avenue NW, Washington, DC 20420
- Jennifer Moser, Ph.D., Associate Director, Scientific Programs
  US Department of Veterans Affairs, 810 Vermont Avenue NW, Washington, DC 20420
- Jennifer E. Deen, B.S., Associate Director, Cohort & Public Relations
  US Department of Veterans Affairs, 810 Vermont Avenue NW, Washington, DC 20420

**MVP Steering Committee**
- Co-Chair: J. Michael Gaziano, M.D., M.P.H.
  VA Boston Healthcare System, 150 S. Huntington Avenue, Boston, MA 021
- Co-Chair: Dave Oslin, M.D.
  Philadelphia VA Medical Center, 3900 Woodland Avenue, Philadelphia, PA 19104
- Sumitra Muralidhar, Ph.D., Ex-Officio
  US Department of Veterans Affairs, 810 Vermont Avenue NW, Washington, DC 20420
- Drew Helmer, M.D., M.S.
  Michael E. DeBakey VA Medical Center, 2002 Holcombe Boulevard, Houston, TX 77030
- Adriana Hung, M.D., M.P.H.
  VA Tennessee Valley Healthcare System, 1310 24th Avenue, South Nashville, TN 37212
- Philip S. Tsao, Ph.D.
  VA Palo Alto Health Care System, 3801 Miranda Avenue, Palo Alto, CA 94304
- Deepak Voora, M.D.
  Durham VA Medical Center, 508 Fulton Street, Durham, NC 27705

**MVP Co-Principal Investigators**
- J. Michael Gaziano, M.D., M.P.H.
  VA Boston Healthcare System, 150 S. Huntington Avenue, Boston, MA 02130
- Philip S. Tsao, Ph.D.
  VA Palo Alto Health Care System, 3801 Miranda Avenue, Palo Alto, CA 94304

**MVP Core Operations**
- Jessica V. Brewer, M.P.H., Director, MVP Cohort Operations
  VA Boston Healthcare System, 150 S. Huntington Avenue, Boston, MA 02130
- Kelly Cho, M.P.H, Ph.D., Director, MVP Phenomics
  VA Boston Healthcare System, 150 S. Huntington Avenue, Boston, MA 02130
- Lori Churby, B.S., Director, MVP Regulatory Affairs
  VA Palo Alto Health Care System, 3801 Miranda Avenue, Palo Alto, CA 94304
- Yonghui Jia, Ph.D., Director, VA Central Biorepository
  VA Boston Healthcare System, 150 S. Huntington Avenue, Boston, MA 02130
- Jacob T. Kean, Ph.D., Acting Director, VA Informatics and Computing Infrastructure (VINCI)
  VA Salt Lake City Health Care System, 500 Foothill Drive, Salt Lake City, UT 84148
- Saiju Pyarajan Ph.D., Director, Data and Computational Sciences
  VA Boston Healthcare System, 150 S. Huntington Avenue, Boston, MA 02130
- Robert Ringer, Pharm.D., Director, VA Albuquerque Central Biorepository
  New Mexico VA Health Care System, 1501 San Pedro Drive SE, Albuquerque, NM 87108
- Luis E. Selva, Ph.D., Director, MVP Biorepository Coordination
  VA Boston Healthcare System, 150 S. Huntington Avenue, Boston, MA 02130

- Shahpoor (Alex) Shayan, M.S., Director, MVP PRE Informatics
  VA Boston Healthcare System, 150 S. Huntington Avenue, Boston, MA 02130
- Brady Stephens, M.S., Principal Investigator, MVP Information Center
  Canandaigua VA Medical Center, 400 Fort Hill Avenue, Canandaigua, NY 14424
- Stacey B. Whitbourne, Ph.D., Director, MVP Cohort Development and Management
  VA Boston Healthcare System, 150 S. Huntington Avenue, Boston, MA 02130

### Supplement 1. Formal definitions of causal estimands in MR studies of time-varying exposures and medication use

Let $k = 0, 1, 2, \dots, K$ denote a time interval with $k = 0$ denoting the time of conception. For each individual, $A_k$ represent the exposure at the beginning of interval $k$, $M_k$ an indicator (1: yes, 0: no) for medication use during interval $k$, and $Y_{k+1}$ an indicator (1: yes, 0: no) for the development of the outcome before the start of the interval $k + 1$. To represent history from time 0, we use an overbar—that is, $\bar{A}_k = (A_0, A_1, \dots, A_k)$ represents an individual's exposure history from time 0 until time $k$ and $\bar{M}_k = (M_0, M_1, \dots, M_k)$ represents an individual's history of medication use from time 0 until time $k$. Let $Y_k^g$ represent the outcome that would have been observed (i.e., the counterfactual outcome) at time $k$ had they followed an intervention $g$, where $g$ may represent an intervention on the biomarker alone (e.g., $g \equiv \bar{g}_K = \bar{a}_K$) or a joint intervention on both the biomarker and medication use (e.g., $g \equiv \bar{g}_K = (\bar{a}_K, \bar{m}_K)$).

For simplicity, we discuss causal estimands for the lifetime effect and the lifetime effect under no medication use.

<u>Lifetime effect</u>

Formally, the lifetime effect can be defined as the contrast between (i) the mean counterfactual outcome had everyone received a hypothetical intervention that increases the natural value of the exposure at each time $k$ by a fixed quantity $c$, and (ii) the mean counterfactual outcome had everyone received their observed (or natural) value of the exposure:

$$\mathrm{E}\left[Y_{K+1}^{g=(\bar{A}_K+\bar{c})}\right] - \mathrm{E}\left[Y_{K+1}^{g'=\bar{A}_K}\right]$$

Alternatively, the lifetime effect can be defined as the contrast between (iii) the mean counterfactual outcome under a hypothetical intervention that keeps the exposure at a fixed level $a$ at all times and (iv) the mean counterfactual outcome under a hypothetical that keeps the exposure at a fixed level $a'$ at all times:

$$\mathrm{E}\left[Y_{K+1}^{g=\bar{a}_K}\right] - \mathrm{E}\left[Y_{K+1}^{g'=\bar{a}'_K}\right]$$

<u>Lifetime effect under no medication use</u>

Formally, the lifetime effect under no medication use can be defined as a contrast between (i) the mean counterfactual outcome had everyone received a hypothetical intervention that increases the natural value of the exposure at each time $k$ by a fixed quantity $c$ with concurrent prevention of medication use, and (ii) the mean counterfactual outcome had everyone received their observed (or natural) value of the exposure and prevention of medication use:

$$\mathrm{E}\left[Y_{K+1}^{g=(\bar{A}_K+\bar{c},\bar{m}_K=\bar{0})}\right] - \mathrm{E}\left[Y_{K+1}^{g'=(\bar{A}_K,\bar{m}_K=\bar{0})}\right]$$

Alternatively, the lifetime effect under no medication use can be defined as the contrast between (iii) the mean counterfactual outcome under a hypothetical intervention that maintains the exposure at a fixed level $a$ with concurrent prevention of medication use and (iv) the mean counterfactual outcome under a hypothetical intervention that maintains the exposure at a fixed level $a'$ with concurrent prevention of medication use:

$$\mathrm{E}\left[Y_{K+1}^{g=(\bar{a}_K,\bar{m}_K=\bar{0})}\right] - \mathrm{E}\left[Y_{K+1}^{g'=(\bar{a}'_K,\bar{m}_K=\bar{0})}\right]$$

### Supplement 2: Data generating mechanisms for simulations of LDL cholesterol (LDL-C), statin use and coronary artery disease

To simulate realistic distributions of LDL-C ($A_k$), statin use ($M_k$) and coronary artery disease ($Y_k$), we used the following data generating models for each individual $i$ at each time $k = 0$ until $k = 9$ or until $Y_{i,k+1} = 1$ was generated, whichever came first.

Causal time-fixed instrument

$$Z_i \sim B(2, 0.5)$$

Time-fixed confounder between LDL-C and coronary artery disease

$$U_i \sim \mathcal{N}(0, 1)$$

Time-fixed confounder between statin use and coronary artery disease

$$W_i \sim \mathcal{N}(0, 1)$$

Time-varying LDL-C levels

$$A_{i,k} \sim \psi(\mu_{A,k}, \sigma_{A,k}, 10, \infty)$$

where

- $\psi(\mu, \sigma, a, b)$ is a truncated normal distribution with mean $\mu$, standard deviation $\sigma$, and bounds of $[a, b]$
- $\mu_{A,k} = 130 + 7Z_i + 25U_i + 0.2(A_{i,k-1} - 130) + 0.1(A_{i,k-2} - 130) - 30M_{k-1}$
- $\sigma_{A,k} = 15$ for $k = 0$; $\sigma_{A,k} = 10$ for $k = 1$; and $\sigma_{A,k} = 5$ for $k \geq 2$

Time-varying statin use

$$M_{i,k} \sim \begin{cases} B(1,1) & \text{if } k = 0 \text{ or } M_{i,k-1} = 1 \\ B(1, p_{M,k}) & \text{otherwise} \end{cases}$$

where

- $\text{logit}(p_{M,k}) = \log\left(\frac{0.01}{0.99}\right) + 0.25\left(\frac{A_{i,k}}{20}\right)\mathrm{I}(A_{i,k} > 100) + 1.3W_i$

Indicator for development of coronary artery disease

$$Y_{i,k+1} \sim B(1, p_{Y,k+1})$$

where

- $\text{logit}(p_{Y,k+1}) = \text{logit}(\lambda_{k+1}) + 1.5U_i + 1.25W_i +$
$$(\beta_{AY} + \beta_{AYk}k)\,\frac{A_{i,k} - 130}{39} + \frac{1}{2}(\beta_{AY} + \beta_{AYk}k)\,\frac{A_{i,k-1} - 130}{39} + \frac{1}{3}(\beta_{AY} + \beta_{AYk}k)\,\frac{A_{i,k-2} - 130}{39} + (\beta_{MY} + \beta_{MYk}k)M_k + \frac{1}{2}(\beta_{MY} + \beta_{MYk}k)M_{k-1} + \frac{1}{3}(\beta_{MY} + \beta_{MYk}k)M_{k-2}$$
- $\lambda_{k+1} = \frac{1}{8}f(k|\mu = 5.5, \sigma^2 = 4)$, where $f(x|\mu, \sigma^2)$ is the probability density evaluated at $x$ for a normal distribution with mean $\mu$ and variance $\sigma^2$

To create a scenario in which participants began follow-up at different ages, each simulated participant was randomly assigned an age at start of follow-up drawn from a normal distribution with a mean of 52.5 years and a standard deviation of 6.25. Assigned ages were rounded down to the nearest multiple of five (to correspond with follow-up time points) and truncated to fall within the range of 30 to 75 years.

We simulated the following four scenarios (scenarios 1 and 2 are also described in the main text of the manuscript), and used the following values of $\beta_{AY}$ and $\beta_{MY}$ for each of the scenarios:

| Scenario | Description | Parameters |
|---|---|---|
| 1 | LDL-C had no direct effect on coronary artery disease<br>Statin use had no direct effect on coronary artery disease | $\beta_{AY} = 0$<br>$\beta_{AYk} = 0$<br>$\beta_{MY} = 0$<br>$\beta_{MYk} = 0$ |
| 2 | LDL-C had a direct harmful effect on coronary artery disease<br>Statin use had a direct protective effect on coronary artery disease | $\beta_{AY} = 0.25$<br>$\beta_{AYk} = 0.02$<br>$\beta_{MY} = -0.6$<br>$\beta_{MYk} = -0.06$ |
| 3 | LDL-C had no direct effect on coronary artery disease<br>Statin use had a direct protective effect on coronary artery disease | $\beta_{AY} = 0$<br>$\beta_{AYk} = 0$<br>$\beta_{MY} = -0.6$<br>$\beta_{MYk} = -0.06$ |
| 4 | LDL-C had a direct harmful effect on coronary artery disease<br>Statin use had no direct effect on coronary artery disease | $\beta_{AY} = 0.25$<br>$\beta_{AYk} = 0.02$<br>$\beta_{MY} = 0$<br>$\beta_{MYk} = 0$ |

### Supplement 3: Statistical analysis for simulations of LDL-C, statin use and coronary artery disease

For two-stage least-squares, we used linear regression in the first stage and Poisson regression in the second stage.

For g-estimation of structural nested cumulative failure time models, we fit the following model and blip function:

$$\exp[\gamma_k(\bar{A}_m, Z; \psi)] = \begin{cases} \dfrac{\mathrm{E}\left[Y_k^{(\bar{A}_m, \underline{0})} \middle| \bar{A}_m, Z, Y_m = 0\right]}{\mathrm{E}\left[Y_k^{(\bar{A}_{m-1}, \underline{0})} \middle| \bar{A}_m, Z, Y_m = 0\right]} & \text{if } Y_m = 0 \\ 1 & \text{if } Y_m = 1 \end{cases}$$

where $\exp[\gamma_k(\bar{A}_m, Z; \psi)] = 1 + \frac{\exp(\psi A_m) - 1}{k - m}$.[1]

The IP weights were defined as:

$$W_k^{\bar{M}} = \begin{cases} \displaystyle\prod_{j=0}^{k} \frac{1}{\Pr\left[M_j = 0 \middle| Z, W, \bar{A}_j, \bar{M}_{j-1} = 0, \bar{Y}_j = 0\right]} & \text{if } M_j = 0 \\ 0 & \text{if } M_j = 1 \end{cases}$$

The denominator of the weights were estimated using a pooled logistic regression model that included flexible functional forms for the time-varying intercept, the instrument $Z$, confounder $W$, and past LDL-C measures $\bar{A}_k$:

$$\Pr\left[M_j = 0 \middle| Z, W, \bar{A}_M, \bar{M}_{k-1} = 0, \bar{Y}_k = 0\right] = \beta_{0,k} f(k) + \beta_1 f(Z) + \beta_2 f(W) + \beta_3 f(\bar{A}_k)$$

In the IV analyses using two-staged least squares, each individual was assigned a weight of $W_{k=K}^{\bar{M}}$ since the outcome status is evaluated at time $K$. In the IV analyses using structural nested cumulative failure time models, each individual was assigned a time-varying weight of $W_k^{\bar{M}}$ at time $k$.

Estimates were highly sensitive to the level of truncation for the weights, with greater bias observed as weights were truncated at a lower percentile. However, not truncating the weights risked non-convergence in some simulation iterations. Therefore, we truncated the weights at the 99.999th percentile.

### Supplement 4: Violation of the Instrumental Conditions under Different Statin Use Adjustment Approaches

For scenario 1 (neither LDL-C, $A_k$, nor statin use, $M_k$, has a direct effect on the outcome, $Y_{k+1}$) and scenario 2 (LDL-C, $A_k$, has a direct harmful effect and statin use, $M_k$, has a direct protective effect on the outcome, $Y_{k+1}$), we outline some of the potential violations of the instrumental conditions under different statin adjustment approaches across different MR study designs: one where follow-up begins at $k = 0$ (with all participants are non-users at start of follow-up) and another where follow-up begins at $k = 1$ (when some participants may already be prevalent users). The violations are described via their biasing paths and the anticipated direction of bias.

In the causal diagrams below, the dashed arrows represent arrows that would be absent in scenario 1 but present in scenario 2. In the biasing paths, variables listed in parentheses represent alternative variables that could be included at that step, reflecting different paths that would result in the biasing pathway being opened.

| **Adjustment approach for statin use** | **Start of follow-up** | **Causal diagram** | **Examples of biasing paths and anticipated direction of bias** [a] |
|---|---|---|---|
| Unadjusted | $k = 0$ | 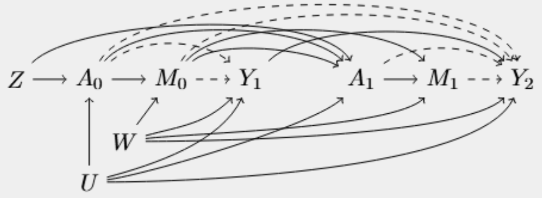 | <u>For scenario 1:</u> none<br><u>For scenario 2:</u> none |
| | $k = 1$ | 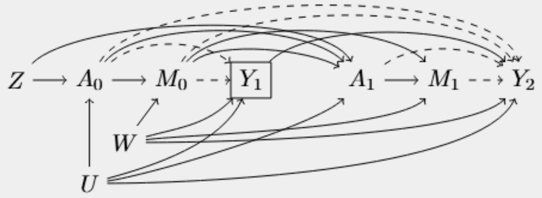 | <u>For scenario 1:</u> none<br><u>For scenario 2 only</u>:<br>$\downarrow: Z \overset{+}{\rightarrow} A_0 \overset{+}{\rightarrow} \boxed{Y_1} \overset{+}{\leftarrow} (W\,;\,U) \overset{+}{\rightarrow} Y_2$<br>$\downarrow: Z \overset{+}{\rightarrow} A_0 \overset{+}{\rightarrow} \boxed{Y_1} \overset{-}{\leftarrow} M_0 \overset{-}{\rightarrow} Y_2$<br>$\downarrow: Z \overset{+}{\rightarrow} A_0 \overset{+}{\rightarrow} \boxed{Y_1} \overset{-}{\leftarrow} M_0 \overset{-}{\rightarrow} A_1 \overset{+}{\rightarrow} Y_2$<br>$\downarrow: Z \overset{+}{\rightarrow} A_0 \overset{+}{\rightarrow} \boxed{Y_1} \overset{-}{\leftarrow} M_0 \overset{+}{\rightarrow} M_1 \overset{-}{\rightarrow} Y_2$<br>$\uparrow: Z \overset{+}{\rightarrow} A_0 \overset{+}{\rightarrow} M_0 \overset{-}{\rightarrow} \boxed{Y_1} \overset{+}{\leftarrow} (W; U) \overset{+}{\rightarrow} Y_2$ |

| Adjustment approach for statin use | Start of follow-up | Causal diagram | Examples of biasing paths and anticipated direction of bias [a] |
|---|---|---|---|
| Restricted to non-users at start of follow-up | $k = 0$ | 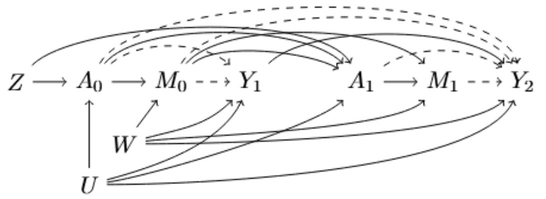 | For scenario 1: none<br>For scenario 2: none |
| | $k = 1$ | 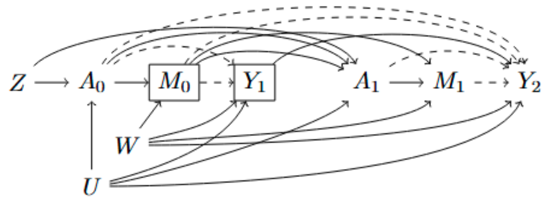 | For both scenarios:<br>$\downarrow: Z \xrightarrow{+} \boxed{A_0} \xleftarrow{+} U \xrightarrow{+} Y_2$<br>$\downarrow: Z \xrightarrow{+} A_0 \xrightarrow{+} \boxed{M_0} \xleftarrow{+} W \xrightarrow{+} Y_2$<br>$\uparrow: Z \xrightarrow{+} \boxed{A_0} \xleftarrow{+} U \xrightarrow{+} \boxed{Y_1} \xleftarrow{+} W \xrightarrow{+} Y_2$<br>For scenario 2 only:<br>$\downarrow: Z \xrightarrow{+} A_0 \xrightarrow{+} \boxed{Y_1} \xleftarrow{+} (W; U) \xrightarrow{+} Y_2$<br>$\downarrow: Z \xrightarrow{+} \boxed{A_0} \xleftarrow{+} U \xrightarrow{+} A_1 \xrightarrow{+} Y_2$<br>$\uparrow: Z \xrightarrow{+} A_0 \xrightarrow{+} \boxed{M_0} \xleftarrow{+} W \xrightarrow{+} M_1 \xrightarrow{-} Y_2$<br>$\uparrow: Z \xrightarrow{+} \boxed{A_0} \xleftarrow{+} U \xrightarrow{+} A_1 \xrightarrow{+} M_1 \xrightarrow{-} Y_2$ |
| Restricted to non-users throughout follow-up | $k = 0$ | 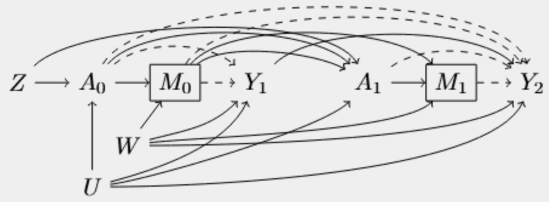 | For both scenarios:<br>$\downarrow: Z \xrightarrow{+} \boxed{(A_0; A_1)} \xleftarrow{+} U \xrightarrow{+} (Y_1; Y_2)$<br>$\downarrow: Z \xrightarrow{+} A_0 \xrightarrow{+} \boxed{M_0} \xleftarrow{+} W \xrightarrow{+} (Y_1; Y_2)$<br>$\downarrow: Z \xrightarrow{+} A_1 \xrightarrow{+} \boxed{M_1} \xleftarrow{+} W \xrightarrow{+} (Y_1; Y_2)$<br>$\uparrow: Z \xrightarrow{+} A_0 \xrightarrow{+} \boxed{M_0} \xleftarrow{+} W \xrightarrow{+} \boxed{M_1} \xleftarrow{+} A_1 \xleftarrow{+} U \xrightarrow{+} (Y_1; Y_2)$<br>$\uparrow: Z \xrightarrow{+} A_1 \xrightarrow{+} \boxed{M_1} \xleftarrow{+} W \xrightarrow{+} \boxed{M_0} \xleftarrow{+} A_0 \xleftarrow{+} U \xrightarrow{+} (Y_1; Y_2)$<br>For scenario 2 only:<br>$\uparrow: Z \xrightarrow{+} A_0 \xrightarrow{+} \boxed{M_0} \xleftarrow{+} W \xrightarrow{+} \boxed{M_1} \xleftarrow{+} A_1 \xrightarrow{+} Y_2$<br>$\uparrow: Z \xrightarrow{+} A_1 \xrightarrow{+} \boxed{M_1} \xleftarrow{+} W \xrightarrow{+} \boxed{M_0} \xleftarrow{+} A_0 \xrightarrow{+} (Y_1; Y_2)$ |

| Adjustment approach for statin use | Start of follow-up | Causal diagram | Examples of biasing paths and anticipated direction of bias [a] |
|---|---|---|---|
| | $k = 1$ | 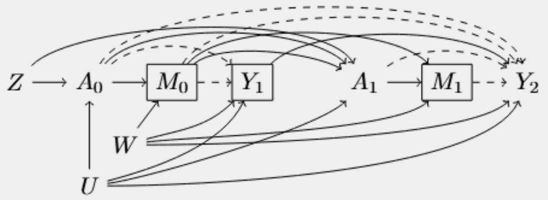 | For both scenarios: same as those described above (for $k = 0$) but only for $Y_2$ as the outcome, plus<br>$\uparrow: Z \xrightarrow{+} \boxed{A_0} \xleftarrow{+} U \xrightarrow{+} \boxed{Y_1} \xleftarrow{+} W \xrightarrow{+} Y_2$<br>For scenario 2 only: same as those described above (for $k = 0$) but only for $Y_2$ as the outcome, plus<br>$\downarrow: Z \xrightarrow{+} A_0 \xrightarrow{+} \boxed{Y_0} \xleftarrow{+} (W;U) \xrightarrow{+} Y_2$<br>$\downarrow: Z \xrightarrow{+} A_0 \xrightarrow{+} \boxed{Y_0} \xleftarrow{+} U \xrightarrow{+} A_1 \xrightarrow{+} Y_2$<br>$\uparrow: Z \xrightarrow{+} A_0 \xrightarrow{+} \boxed{Y_0} \xleftarrow{+} W \xrightarrow{+} \boxed{M_1} \xleftarrow{+} A_1 \xrightarrow{+} Y_2$<br>$\uparrow: Z \xrightarrow{+} A_1 \xrightarrow{+} \boxed{M_1} \xleftarrow{+} W \xrightarrow{+} \boxed{M_0} \xleftarrow{+} A_0 \xrightarrow{+} Y_2$ |
| IP weighting among non-users at start of follow-up | $k = 0$ | 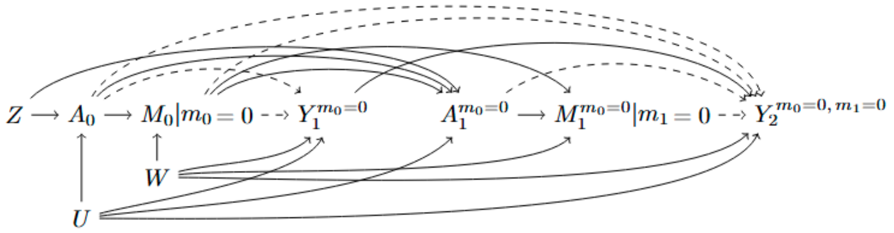 | For scenario 1: none<br>For scenario 4: none |
| | $k = 1$ | 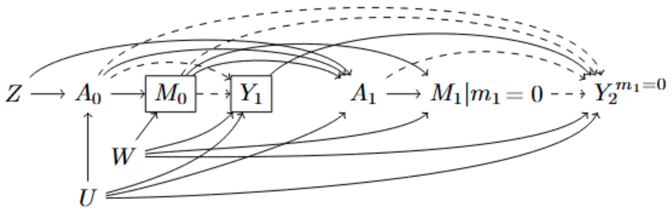 | For both scenarios:<br>$\downarrow: Z \xrightarrow{+} \boxed{A_0} \xleftarrow{+} U \xrightarrow{+} Y_2$<br>$\downarrow: Z \xrightarrow{+} A_0 \xrightarrow{+} \boxed{M_0} \xleftarrow{+} W \xrightarrow{+} Y_2$<br>$\uparrow: Z \xrightarrow{+} \boxed{A_0} \xleftarrow{+} U \xrightarrow{+} \boxed{Y_1} \xleftarrow{+} W \xrightarrow{+} Y_2$<br>For scenario 2 only:<br>$\downarrow: Z \xrightarrow{+} A_0 \xrightarrow{+} \boxed{Y_1} \xleftarrow{+} (W;U) \xrightarrow{+} Y_2$<br>$\downarrow: Z \xrightarrow{+} \boxed{A_0} \xleftarrow{+} U \xrightarrow{+} A_1 \xrightarrow{+} Y_2$ |

[a] A plus (+) symbol over an arrow indicates a positive association. A minus (-) symbol over an arrow indicates an inverse association. The overall direction of bias from the biasing path is indicated using a down arrow (↓) for downward bias and an up arrow (↑) for upward bias. This is determined by multiplying the positive and negative signs associated with each arrow in the pathway, with the sign being reversed whenever a variable is conditioned upon.

In general, biasing paths are only absent when follow-up begins at $k = 0$ and no variables are conditioned on, such as in an unadjusted analysis, when restricting to non-users at baseline if no participants are using statins at that point, or when applying IP weighting. Another exception is an unadjusted analysis starting at $k = 0$ in a scenario where both LDL-C and statin use have no effect on the outcome (i.e., scenario 1).

All other scenarios include a combination of upward and downward biases. The number of open biasing paths increases as more variables are conditioned upon—for example, with a later start of follow-up (i.e., conditioning on more $Y_k$ variables) or when conditioning on statin use ($M_k$) across multiple time points. Furthermore, scenario 2 introduces even more open biasing paths compared to scenario 1, due to the presence of direct effects from $A_k$ and $M_k$ to $Y_{k+1}$. In our simulations, both LDL-C and statin use were modeled to have strong effects on the outcome, so biasing paths unique to scenario 2 likely introduced greater bias than those present in both scenarios. This may explain why the direction of bias shifted between scenarios when, for example, analyses were conditioned on non-users at baseline and throughout follow-up; biasing paths specific to scenario 2 introduced an upward bias, altering the overall direction of bias.

Despite these patterns, it remains difficult, even in simulations, to predict the precise direction and magnitude of bias, giving the complex interplay of multiple opposing biasing paths. The accompanying table simplifies these ideas by considering these biases for two time points. In contrast, our simulations incorporated 10 time points, which greatly increases the number and complexity of potential biasing paths. In real-world data, with many additional variables (e.g., multiple and possibly time-varying confounders $U$ and $W$), predicting bias becomes even more challenging.

### Supplementary Tables

**Table S1. Variables used in the Mendelian randomization study of LDL-C and coronary artery disease using observational data from the Million Veteran Program (January 1, 2002 to December 31, 2020)**

| Variable | Variable type | Detail | Codes |
|---|---|---|---|
| Age | Continuous (years) | Based on records in the MVP Roster file. | N/A |
| Sex | Binary: Male / Female | Based on records in the MVP Core Demographics file. | N/A |
| Harmonized ancestry and race/ethnicity (HARE) | Categorical: Non-Hispanic white / Non-Hispanic Black / Non-Hispanic Asian / Hispanic | Genetically defined ancestry based on the use of a supervised machine learning algorithm applied to self-identified racial and ethnic (SIRE) background | N/A |
| Primary care visit in the past year | Binary: Yes / No | Time-updated count of in-person or telehealth primary care visit, based on stop codes in any position. Based on records in the *Outpatient* domain. | **Stop codes**: 156, 157, 170-178, 322, 323, 338, 341, 342, 348, 350, 531, 534, 539, 704 |
| Hospitalization in the past 12 months | Binary: Yes / No | Receipt of inpatient care. Based on records in the *Inpatient* domain. | N/A |
| Smoking status | Categorical: Never / Current / Former | Time-updated measure of most recent self-reported smoking status. Based on records in the *Health Factors* domain. | N/A |
| Body mass index | Continuous ($kg/m^2$) | Time-updated measure calculated using the most recent height and weight measurements. Based on records in the *Vital* domain. | N/A |
| Systolic blood pressure | Continuous (mmHg) | Time-updated measure based on records in the *Vital* domain. | N/A |

| Variable | Variable type | Detail | Codes |
|---|---|---|---|
| Diastolic blood pressure | Continuous (mmHg) | Time-updated measure based on records in the *Vital* domain. | N/A |
| Total cholesterol | Continuous (mg/dL) | Time-updated measure of total cholesterol identified using an adjudicated list. Based on records in the *Labs* domain. | **Short Name**: TotChol |
| Triglycerides | Continuous (mg/dL) | Time-updated measure of triglycerides identified using an adjudicated list. Based on records in the *Labs* domain. | **Short Name**: Trig |
| High-density lipoprotein (HDL) cholesterol | Continuous (mg/dL) | Time-updated measure of HDL cholesterol identified using an adjudicated list. Based on records in the *Labs* domain. | **Short Name**: HDLC |
| Low-density lipoprotein (LDL) cholesterol | Continuous (mg/dL) | Time-updated measure of LDL-C identified using an adjudicated list. Based on records in the *Labs* domain. | **Short Name**: LDLC |
| Alanine transaminase (ALT) | Continuous (IU/L) | Time-updated measure of ALT identified using an adjudicated list. Based on records in the *Labs* domain. | **Short Name**: ALT |
| Aspartate aminotransferase (AST) | Continuous (IU/L) | Time-updated measure of AST identified using an adjudicated list. Based on records in the *Labs* domain. | **Short Name**: AST |
| Use of a lipid-lowering therapy | Binary: Yes / No | Lipid-lowering therapies, identified using an adjudicated list. Based on records in the *Medications* domain. | **VA Classification**: CV350<br><br>Includes the following generic drug names: colestipol, dextrophyroxine, evolocumab, ezetimibe, ezetimibe/simvastatin, fenofibrate, fenofibric acid, fish oil, gemfibrozil, icosapent ethyl, lomitapide, mevacor, mipomersen niacinamide, omega-3, omega-3 acid, probucol, rosuvastatin, statin, alirocumab, atorvastatin, atromid, bezafibrate, bezalip retard, cholestyramine, choloxin, |

| Variable | Variable type | Detail | Codes |
|---|---|---|---|
| | | | clofibrate, colesevelam. Based on records in the *Medications* domain. |
| Use of an antianginal therapy | Binary: Yes / No | Antianginal therapies, identified using an adjudicated list. Based on records in the *Medications* domain. | **VA Classification**: CV250<br><br>Includes the following generic drug names: erythrityl tetranitrate, glyceryl trinitrate, isosorbide, isosorbide dinitrate, isosorbide mononitrate, meprobamate pentaerythritol, nitroglycerin, nitroglycerin/petrolatum, nitropropane/petrolatum, pentaerythritol, pentaerythritol tetranitrate, ranolazine. |
| Use of an anticoagulant therapy | Binary: Yes / No | Anticoagulant therapy, identified using an adjudicated list. Based on records in the *Medications* domain. | **VA Classification**: BL110<br><br>Includes the following generic drug names: acenocoumarol, anisindione, apixaban, argatroban, betrixaban, bivalirudin, certoparin sodium, citrate, citric acid/sodium citrate/sodium phosphate/dextrose, dabigatran, dalteparin, danaparoid, dicumarol, dihydroergotamine/heparin, dihydroergotamine/heparin/lidocaine, edoxaban, enoxaparin, eptifibatide, fondaparinux, fraxiparine, heparin, heparin/dextrose, heparin/sodium chloride, lepirudin, rivaroxaban, sodium citrate, tibrofiban, tinzaparin, warfarin, ximelagatran. |
| Use of an antihypertensive therapy | Binary: Yes / No | Antihypertensive therapy, identified using an adjudicated list. Based on records in the *Medications* domain. | **VA Classification**: CV400, CV490<br><br>Includes the following generic drug names: acebutolol, atenolol, betaxolol, bisoprolol, bisoprolol/hydrochlorothiazide, carteolol, carvedilol, esmolol, labetalol, metoprolol, metoprolol succinate, metoprolol tartrate, nadolol, nebivolol, penbutolol, pindolol, propranolol, sotalol, timolol, aliskiren/amlodipine/hydrochlorothiazide, aliskiren/hydrochlorothiazide, aliskiren/valsartan, amiloride/hydrochlorothiazide, amlodipine/benazepril, amlodipine/hydrochlorothiazide/olmesartan, amlodipine/hydrochlorothiazide/valsartan, amlodipine/olmesartan, amlodipine/valsartan, |

| Variable | Variable type | Detail | Codes |
|---|---|---|---|
| | | | atenolol/chlorthalidone, azilsartan/chlorthalidone, benazepril/hydrochlorothiazide, bendroflumethiazide/nadolol, bendroflumethiazide/rauwolfia, bisoprolol/hydrochlorothiazide, candesartan/hydrochlorothiazide, captopril/hydrochlorothiazide, chlorothiazide/methyldopa, chlorothiazide/reserpine, chlorthalidone/clonidine, chlorthalidone/reserpine, clonidine/chlorthalidone, deserpidine/hydrochlorothiazide, deserpidine/methyclothiazide, deserpidine/pargyline, enalapril/felodipine, enalapril/hydrochlorothiazide, fosinopril/hydrochlorothiazide, guanethidine/hydrochlorothiazide, hydralazine, hydralazine/hydrochlorothiazide, hydralazine/hydrochlorothiazide/reserpine, hydralazine/isosorbide, hydralazine/reserpine, hydrochlorothiazide/enalapril, hydrochlorothiazide/hydralazine, hydrochlorothiazide/hydrochlorothiazide/reserpine, hydrochlorothiazide/irbesartan, hydrochlorothiazide/lisinopril, hydrochlorothiazide/losartan, hydrochlorothiazide/methyldopa, hydrochlorothiazide/metoprolol, hydrochlorothiazide/moexipril, hydrochlorothiazide/olmesartan, hydrochlorothiazide/propranolol, hydrochlorothiazide/quinapril, hydrochlorothiazide/reserpine, hydrochlorothiazide/telmisartan, hydrochlorothiazide/timolol, hydrochlorothiazide/triamterene, hydrochlorothiazide/valsartan, hydroflumethiazide/reserpine, lisinopril/hydrochlorothiazide, losartan/hydrochlorothiazide, methyclothiazide/pargyline, |

| Variable | Variable type | Detail | Codes |
|---|---|---|---|
| | | | methyldopa/chlorothiazide, methyldopa/hydrochlorothiazide, moexipril/hydrochlorothiazide, nadolol/bendroflumethiazide, nebivolol/valsartan, polythiazide/prazosin, polythiazide/reserpine, prazosin/polythiazide, propranolol/hydrochlorothiazide, quinapril/hydrochlorothiazide, quinethazone, quinethazone/reserpine, rauwolfia/bendroflumethiazide, rauwolfia/flumethiazide/potassium, rescinnamine, reserpine/chlorthalidone, reserpine/hydralazine, reserpine/hydralazine/hydrochlorothiazide, reserpine/hydrochlorothiazide, reserpine/trichlormethiazide, statin, telmisartan/hydrochlorothiazide, trandolapril/verapamil, trichlormethiazide/reserpine, valsartan/hydrochlorothiazide. |
| Use of a beta blocker | Binary: Yes / No | Beta blockers, identified using an adjudicated list. Based on records in the *Medications* domain. | **VA Classification**: CV100<br><br>Includes the following generic drug names: acebutolol, atenolol, betaxolol, bisoprolol, bisoprolol/hydrochlorothiazide, carteolol, carvedilol, esmolol, labetalol, metoprolol, metoprolol succinate, metoprolol tartrate, nadolol, nebivolol, penbutolol, pindolol, propranolol, sotalol, timolol. |
| Coronary artery disease | Binary: Yes / No | Defined as ≥1 diagnosis or procedure code in the *Inpatient* domain or ≥2 diagnosis codes in the *Outpatient* domain | **ICD-9**: 41[0-2].%, 414.%, 429.2, 429.5, 429.7%, V45.82<br>**ICD-10**: I20.0, I2[1-5].% (excluding I25.112 and I25.7[0-9]2), I51.[0-1]%, Z95.5%, Z98.61<br>**ICD-9 Procedure**: 36.01, 36.02, 36.05, 36.1%, 00.66<br>**ICD-10 Procedure**: 021008W, 021[0-1]09[CFW], 021[0-2]0A[389CW], 02100J[389W], 0200K[389W], 021[0-1]0Z%, 021[0-2]49W, 02104A[89W], 02104J3, 02104K[9W], 02104Z[389], 02110J[9W], 02110KW, 02114AW, 02114J9, 02114K3, 02114Z[39], 021209[CW], 021[2-3]0J[389W], 02120K[39W], 02120Z[389C], 02124A9, 02124Z3, 021309W, 02130A[89W], 02130KW, 02130Z[39], 021349C, 02134KW, 02134Z3, 02703[4567DEFGTZ]Z, 02703[DZ]6, 02704[4567DEFGZ]Z, 02704[DZ]6, |

| Variable | Variable type | Detail | Codes |
|---|---|---|---|
| | | | 02713[4567DEFGZ]Z, 02713[DFGZ]6, 02714[4567DEGZ]Z, 02723[4567DEFGZ]Z, 02723[DFGZ]6, 02724[4567DEFZ]Z, 02733[4567DGZ]Z, 02733[GZ]6, 02734[4567DGZ]Z<br>**CPT**: 33150-33536, 929[2-4]% , 92974, 92975 |
| Myopathy | Binary:<br>Yes / No | Defined as ≥1 diagnosis in the *Inpatient* domain or ≥2 diagnoses in the *Outpatient* domain | **ICD-9**: 359.[4-9]%<br>**ICD-10**: G71.3, G71.[8-9]%, G72.[0-2]%, G72.4%, G72.[8-9]%, G73.7, M05.4[0-7]%, M05.49, M33.[0-2]2, M33.92, M34.82, M35.03, M60.%, M62.9 |
| Chronic liver disease | Binary:<br>Yes / No | Defined as ≥1 diagnosis in the *Inpatient* domain or ≥2 diagnoses in the *Outpatient* domain | **ICD-9**: 571.%, 572.%, 573.3%, 070.%<br>**ICD-10**: K70.%, K7[2-5].% |
| Chronic kidney disease or end renal disease | Binary:<br>Yes / No | Defined as ≥1 diagnosis in the *Inpatient* domain or ≥2 diagnoses in the *Outpatient* domain | **ICD-9**: 016.0%, 095.4, 189.0, 189.9, 223.0, 236.91, 249.4%, 250.4%, 271.4, 274.10, 283.11, 403.[0-1]%, 403.91%, 404.0[2-3]%, 404.1[2-3]%, 404.9[2-3]%, 440.1, 442.1, 572.4, 580[0-8].%, 581.%, 591.%, 573.1[2-9]%, 573.2%, 794.4<br>**ICD-10**: A18.11, A52.75, B52.0, C64.%, C68.9, D30.0%, D41.[0-2]%, D59.3, E08.2%, E08.65, E09.2%, E10.2%, E10.65, E11.2%, E11.65, E13.2%, E74.8, I1[2-3].%, I70.1, I72.2, K76.7, M10.3%, M32.1[4-5], M35.04, N0.%, N13.[1-3]%, N14.%, N15.0, N15.[8-9], N1[6-9], N25.%, N26.1, N26.9, Q61.02, Q61.[1-5]%, Q61.8, Q62.[0-3]%, R94.4<br>**ICD-9 Procedure**: 38.95, 39.93, 39.95, 54.98%, 99.78<br>**ICD-10 Procedure**: 031[3-8]0JD, 03190JF, 031[ABC]0JF, 3E1M39Z, 5A1D[7-9]0Z, 6ABT0BZ |
| Vascular-related diseases | Binary: Yes / No | Includes stroke, angina, acute coronary syndrome, peripheral vascular disease, claudication, cardiomyopathy, heart failure, chronic ischemic heart disease, and deep vein thrombosis. Defined as ≥1 diagnosis in the *Inpatient* domain or ≥2 diagnoses in the *Outpatient* domain | **ICD-9**: 433.[0-9]1, 436%, 437.0, 437.6, 434% (excluding 434.[0-9]0), 413%, 411.1%, 433%, 440.21%, 425%, 428%, 414%, 453.[4-5]%, 453.[7-8]%<br>**ICD-10**: I63.[0-9][0-9]9, I63.[2-5]0, I63.22, I63.59, I67.2, I67.6, I67.89, I20%, I25.7%, I73%, I79.8%, I79.1%, I70.[2-7]1%, I51.81, I4[2-3]%, I50%, I25%, I82.2%, I82.29%, I82.[4-5]%, I82.62%, I82.72%, I82.A%, I82.B%, I82.C% |

| Variable | Variable type | Detail | Codes |
|---|---|---|---|
| Diabetes | Binary: Yes / No | Defined as ≥1 diagnosis in the *Inpatient* domain or ≥2 diagnoses in the *Outpatient* domain | **ICD-9**: 250%<br>**ICD-10**: E1[0-4].[0-9], |
| Chronic obstructive pulmonary disease | Binary: Yes / No | Defined as ≥1 diagnosis in the *Inpatient* domain or ≥2 diagnoses in the *Outpatient* domain | **ICD-9**: 115%, 49[0-1]%, 492.8, 494%, 496%, 748.61, V81.3<br>**ICD-10**: J4[0-3]%, J44[0-1]%, J44.9%, J47% |
| Depression | Binary: Yes / No | Defined as ≥1 diagnosis in the *Inpatient* domain or ≥2 diagnoses in the *Outpatient* domain | **ICD-9**: 296.[2-3]%, 300.4%, 311%<br>**ICD-10**: F32%, F33%, F34.1% |
| Hypothyroidism | Binary: Yes / No | Defined as ≥1 diagnosis in the *Inpatient* domain or ≥2 diagnoses in the *Outpatient* domain | **ICD-9**: 24[3-4]%<br>**ICD-10**: E01.8%, E02%, E03.[2-3]%, E03.[8-9]%, E89.0% |
| Osteoporosis | Binary: Yes / No | Defined as ≥1 diagnosis in the *Inpatient* domain or ≥2 diagnoses in the *Outpatient* domain | **ICD-9**: 733.0%<br>**ICD-10**: M81.0%, M81.6%, M81.8% |
| Loss to follow-up | Binary: Yes / No | Defined as two years after the last LDL-C measurement | N/A |

**Table S2. Summary statistics for the associations between the proposed instruments for LDL-C (based on genetic variants in the *HMGCR*, *PCSK9* and *LDLR* genes) and LDL-C, obtained from the Global Lipids Genetics Consortium**

| Gene | SNP | Position | Reference allele | Alternate allele | Beta | SE | Reference allele frequency |
|---|---|---|---|---|---|---|---|
| *PCSK9* | rs2479394 | chr1:55258652 | G | A | 0.0386 | 0.0041 | 0.2850 |
| *PCSK9* | rs11206510 | chr1:55268627 | T | C | 0.0831 | 0.0050 | 0.8456 |
| *PCSK9* | rs2149041 | chr1:55274725 | G | C | 0.0636 | 0.0049 | 0.1609 |
| *PCSK9* | rs10888897 | chr1:55285649 | C | T | 0.0507 | 0.0042 | 0.6055 |
| *PCSK9* | rs7552841 | chr1:55291340 | T | C | 0.0368 | 0.0044 | 0.3654 |
| *PCSK9* | rs562556 | chr1:55296825 | A | G | 0.0640 | 0.0066 | 0.8061 |
| *HMGCR* | rs10066707 | chr5:74596335 | A | G | 0.0497 | 0.0054 | 0.4169 |
| *HMGCR* | rs2006760 | chr5:74597785 | G | C | 0.0533 | 0.0076 | 1.0000 |
| *HMGCR* | rs2303152 | chr5:74677463 | A | G | 0.0423 | 0.0064 | 0.1201 |
| *HMGCR* | rs17238484 | chr5:74684252 | T | G | 0.0627 | 0.0062 | 0.2533 |
| *HMGCR* | rs5909 | chr5:74691931 | A | G | 0.0617 | 0.0088 | 0.1016 |
| *HMGCR* | rs12916 | chr5:74692295 | C | T | 0.0733 | 0.0038 | 0.4314 |
| *LDLR* | rs6511720 | chr19:11063306 | G | T | 0.2209 | 0.0061 | 0.9024 |
| *LDLR* | rs688 | chr19:11088602 | T | C | 0.0540 | 0.0037 | 0.4472 |

**Table S3. Number of participants and coronary artery disease events for each MR analysis, stratified by age group (<60, 60 to <70) with risk ratios and 95% confidence intervals corresponding to a change in LDL-C from 100 mg/dL to 139 mg/dL via two-stage least squares and g-estimation of structural nested cumulative failure time models**

| Adjustment approach for lipid-lowering therapy use | Age group | Number of participants | Number of events | Risk ratio (95% CI) corresponding to a change in LDL-C from 100 mg/dL to 139 mg/dL | | Number of bootstrapped samples (out of 500) with successful SNCFTM convergence |
|---|---|---|---|---|---|---|
| | | | | Two-stage least squares (TSLS) | Structural nested cumulative failure time models (SNCFTM) [a] | |
| Unadjusted | All | 327,063 | 63,626 | 1.09 (0.97, 1.22) | 1.06 (0.96, 1.20) | 500 |
| | <60 | 192,239 | 26,670 | 1.20 (1.02, 1.43) | 1.20 (0.97, 1.66) | 498 |
| | 60 to <70 | 167,459 | 40,610 | 1.07 (0.90, 1.26) | 0.98 (0.87, 1.10) | 500 |
| Restricted to never users at start of follow-up | All | 279,216 | 49,387 | 1.13 (1.02, 1.28) | 1.06 (0.94, 1.20) | 500 |
| | <60 | 170,159 | 21,063 | 1.19 (0.99, 1.43) | 1.13 (0.91, 1.45) | 499 |
| | 60 to <70 | 120,873 | 27,966 | 1.14 (0.97, 1.33) | 1.04 (0.92, 1.17) | 500 |
| Restrict to never users over follow-up | All | 128,852 | 19,470 | 1.51 (1.18, 1.96) | 1.11 (0.92, 1.94) | 497 |
| | <60 | 82,747 | 7,356 | 1.79 (1.23, 2.96) | 1.24 (0.91, 4.68) | 462 |
| | 60 to <70 | 49,429 | 11,396 | 1.37 (1.02, 1.88) | 1.04 (0.81, 1.28) | 490 |
| IP weighting with restriction to new users at start of follow-up | All | 279,216 | 49,387 | 1.34 (1.07, 1.69) | 1.08 (0.90, 1.34) | 500 |
| | <60 | 170,159 | 21,063 | 1.83 (1.16, 3.11) | 1.22 (0.83, 1.97) | 470 |
| | 60 to <70 | 120,873 | 27,966 | 1.20 (0.92, 1.60) | 1.11 (0.90, 1.70) | 492 |

[a] Confidence intervals were computed as the 2.5th and 97.5th percentiles of the bootstrapped estimates from samples in which the model successfully converged.

### Supplementary Figures

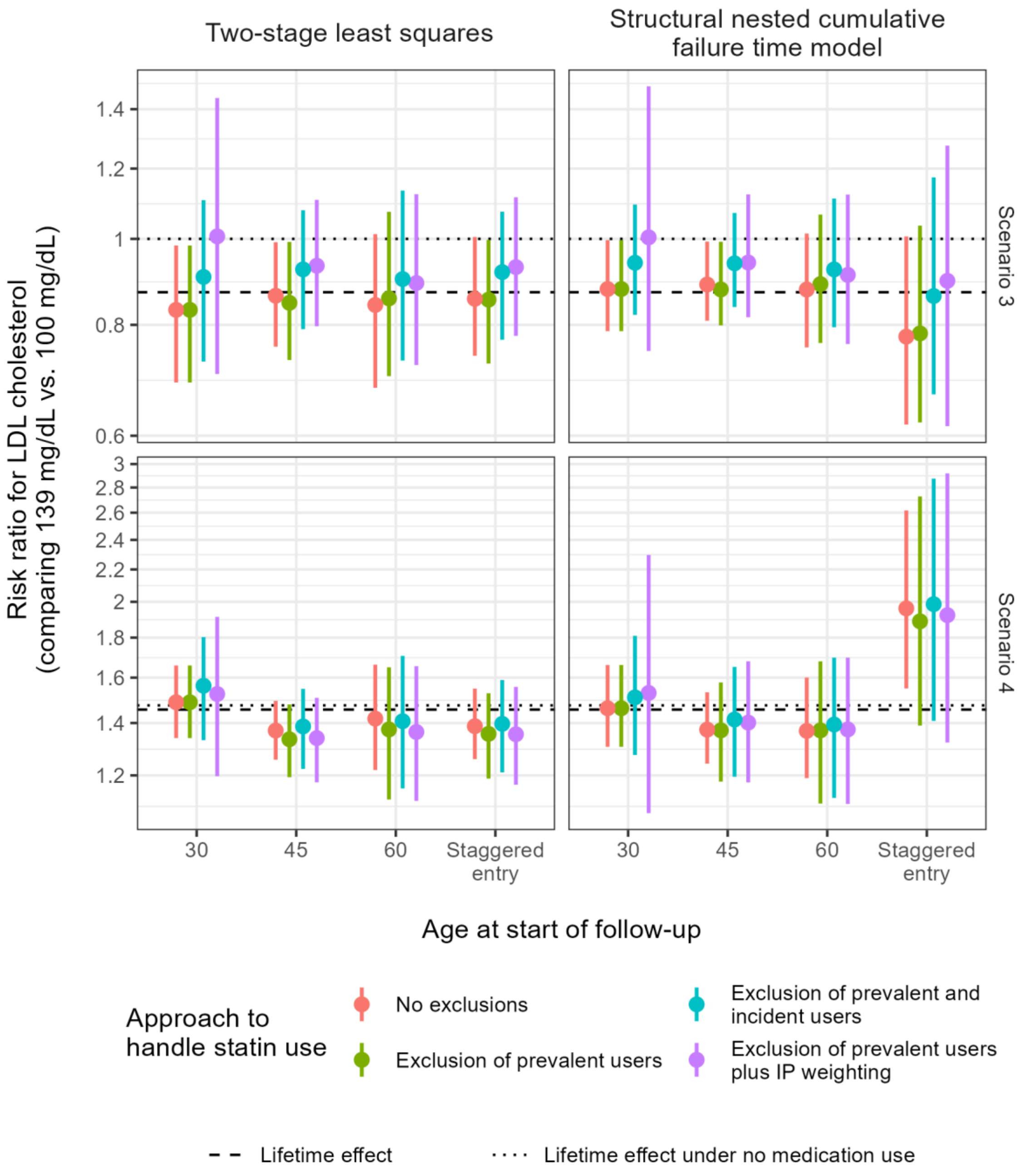

**Figure S1. Risk ratio estimates under two IV methods, summarized across 500 iterations (by mean and 2.5th and 97.5th percentiles). In scenario 3, the effect of LDL-C was null while statins had a direct protective effect. In scenario 4, LDL-C had a direct harmful effect, while the effect of statins was null.**

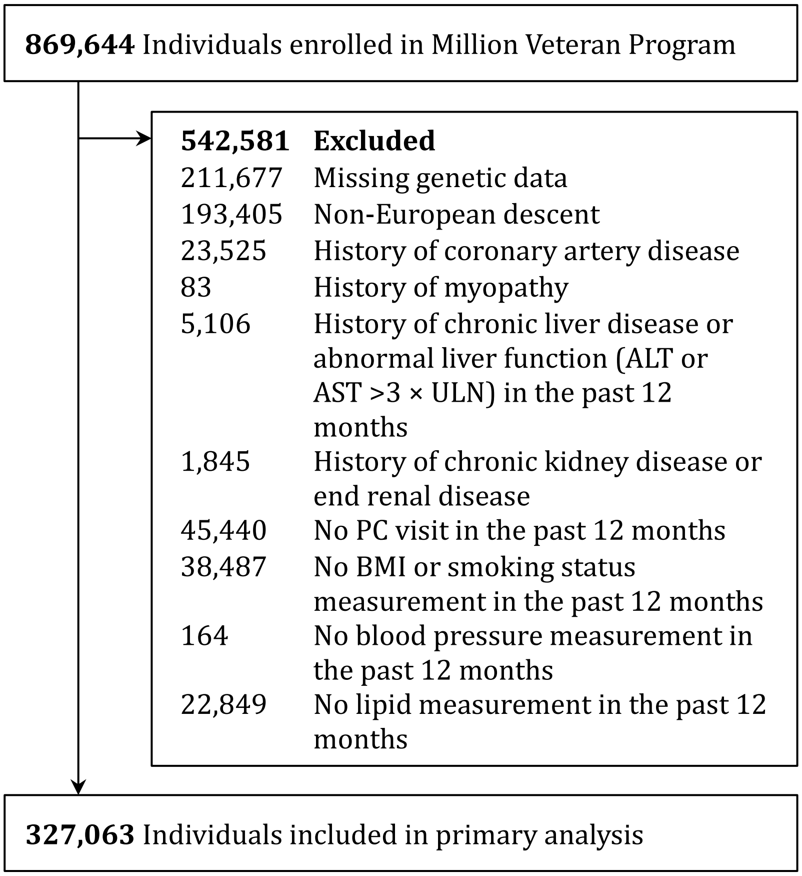

**Figure S2. Selection of eligible individuals for an MR analysis of LDL-C and coronary artery disease using data from the Million Veteran Program**
